# Interpretability as stability under perturbation reveals systematic inconsistencies in feature attribution

**DOI:** 10.64898/2026.04.20.26351354

**Authors:** Natalia Piórkowska, Agnieszka Olejnik, Alan Ostromęcki, Wiktor Kuliczkowski, Andrzej Mysiak, Iwona Bil-Lula

## Abstract

Interpreting machine learning models typically relies on feature attribution methods that quantify the contribution of individual variables to model predictions. However, it remains unclear whether attribution magnitude reflects the true functional importance of features for model performance.

Here, we present a unified interpretability framework integrating permutation-based attribution, feature ablation, and stability under perturbation across multiple feature spaces. Using nested cross-validation and permutation-based null diagnostics, we systematically evaluate the relationship between attribution magnitude and functional dependence in clinical and biomarker-based prediction models.

Attribution magnitude is frequently misaligned with functional importance, with weak to strong negative correlations observed across feature spaces (Spearman ρ ranging from −0.374 to −0.917). Features with high attribution often have limited impact on model performance when removed, whereas features with low attribution can be essential for maintaining predictive accuracy. These discrepancies define distinct classes of interpretability failure, including attribution excess and latent dependence.

Interpretability further depends on feature space composition, and stable, functionally relevant features are not necessarily those with the highest attribution scores. By integrating attribution, functional impact, and stability into a composite Feature Reliability Score, we identify features that remain informative across perturbations and analytical contexts.

These findings indicate that interpretability does not arise from attribution magnitude alone but is better characterized from stability under perturbation. This framework provides a basis for more robust model interpretation and highlights limitations of attribution-centric approaches in high-dimensional and correlated data settings.

## Introduction

The rapid adoption of machine learning in biomedical and clinical research has intensified the need for interpretable and reliable predictive models that can support scientific inference and decision-making [1–3]. In this context, feature attribution methods—such as permutation importance, SHapley Additive exPlanations (SHAP) values, and related approaches—are widely used to quantify the contribution of individual variables to model predictions and to guide feature selection and hypothesis generation [4–6]. These methods are often implicitly treated as proxies for feature relevance.

However, a fundamental assumption underlying this practice has not been rigorously evaluated: that attribution magnitude reflects the true functional importance of a feature for model performance. In reality, attribution methods quantify the sensitivity of model outputs to perturbations in feature distributions, rather than the structural dependence of the model on those features [7,8]. Consequently, high attribution scores may arise from correlations, redundancy, or interaction effects, rather than from direct contribution to predictive performance [9,10].

Recent advances in explainable artificial intelligence have highlighted the instability and context dependence of attribution methods, particularly in high-dimensional and correlated datasets [11–13,18–21]. Attribution values can vary substantially across model configurations, data partitions, and feature representations, raising concerns about their reliability. At the same time, alternative approaches based on structural perturbation—such as feature ablation or leave-one-out retraining—provide complementary insights by directly quantifying the effect of feature removal on model performance [14,15]. Despite their conceptual complementarity, these two perspectives—distributional perturbation and structural perturbation—are rarely integrated within a unified interpretability framework.

This limitation is especially critical in clinical and biomarker-based modeling, where feature spaces often exhibit high redundancy, surrogate measurements, and complex dependency structures [16,17]. In such settings, reliance on attribution magnitude alone may lead to misleading conclusions about feature importance, potentially obscuring clinically or biologically relevant signals.

Here, we address this gap by introducing a unified interpretability framework that integrates permutation-based attribution, feature ablation, stability across model variants, and permutation-based model diagnostics. We systematically evaluate the relationship between attribution magnitude and functional importance across three distinct feature spaces: a combined clinical– biomarker set (FULL), a clinical subset (CLINICAL), and a biomarker-only subset (BIOMARKERS). By incorporating stability measures and null-model diagnostics, we further assess the robustness of feature relevance under multiple perturbation regimes.

We show that attribution magnitude is frequently misaligned with functional importance and that this misalignment depends strongly on feature space composition. Based on these discrepancies, we define distinct classes of interpretability failure, including attribution excess and latent dependence, and demonstrate that features identified as most important by attribution are not necessarily those most critical for predictive performance. Importantly, we further show that reliable features are characterized by stability under perturbation rather than by attribution magnitude alone.

To our knowledge, this study provides the first systematic integration of attribution, ablation, and cross-context stability into a unified framework for identifying interpretability failure modes in machine learning models. Our results suggest that interpretability should be understood as a multi-dimensional and context-dependent property, rather than as a direct consequence of attribution magnitude.

Together, these findings provide a framework for more robust and reproducible model interpretation and highlight fundamental limitations of attribution-centric approaches in high-dimensional and correlated biomedical data settings.

## Methods

### Study design and analytical framework

This study developed a multi-stage analytical pipeline to investigate the relationship between permutation-based feature attribution, functional importance (defined as performance degradation upon feature removal), and interpretability stability in machine learning models. The workflow integrated model training, permutation-based attribution analysis, feature-removal experiments, model-level permutation diagnostics, and contradiction-focused feature profiling into a unified interpretability framework.

Three feature spaces were analyzed independently:

i. a FULL feature set combining clinical and biomarker features,
ii. a CLINICAL subset restricted to routinely available clinical variables, and
iii. a BIOMARKERS subset consisting of circulating biomarker measurements.

All analyses were performed using harmonized preprocessing, model selection, and evaluation procedures across these feature spaces to enable direct comparison of interpretability profiles under different feature-space definitions.

### Model development and evaluation

Models were trained using a nested cross-validation framework to reduce optimistic bias in both performance estimation and downstream interpretability analysis. The outer loop was used for model evaluation, whereas the inner loop was used for model selection and hyperparameter tuning.

Performance was assessed using:

- area under the receiver operating characteristic curve (ROC AUC),
- Matthews correlation coefficient (MCC).

To assess whether model performance exceeded chance-level expectations, permutation-based null distributions were constructed by repeatedly shuffling outcome labels and retraining models under the same validation framework. For each model, permutation-based distributions of ROC AUC and MCC were used to derive:

- empirical *p*-values,
- standardized effect sizes relative to the null distribution.

A composite model diagnostic reliability score was then derived by combining statistical significance and standardized effect-size measures for both ROC AUC and MCC. In the present framework, this score was treated as a variant-level diagnostic context rather than as a direct feature-level component.

### Permutation-based feature attribution

Feature attribution was quantified using permutation importance computed within the outer cross-validation folds. For each feature, the decrease in model performance (ROC AUC) after random permutation of its values was calculated, yielding a measure of attribution magnitude under distributional perturbation.

Feature-level attribution values were aggregated across folds to obtain:

- mean permutation importance,
- rank ordering of features based on attribution magnitude.

Because raw permutation importance may include values close to zero or slightly negative due to sampling variability, non-negative transformed values were used for downstream relative scoring. Accordingly, all normalized attribution-based quantities in this study should be interpreted as relative within feature space, not as absolute effect sizes.

### Feature removal (ablation) analysis

To quantify functional importance, a feature ablation framework was implemented. For each feature, the model was retrained after removing that feature from the input space, and the resulting change in predictive performance was measured.

Two ablation-based metrics were computed:

- change in cross-validated mean AUC,
- change in out-of-fold (OOF) AUC.

The primary downstream measure was the decrease in mean AUC relative to the baseline model, including all features. Negative ablation deltas, corresponding to cases in which feature removal did not reduce performance, were retained in raw audit tables but clipped to zero for downstream relative scoring. Thus, the ablation-based score reflects positive functional dependence rather than any arbitrary signed change in performance.

Ablation was treated as a complementary measure of structural model dependence, not as absolute ground truth.

### Construction of the unified feature reliability framework

Feature-level interpretability was quantified using a composite Feature Reliability Score (FRS). In the final framework, FRS integrated three feature-level components:

- attribution magnitude, derived from non-negative permutation importance,
- functional impact, derived from ablation-based performance degradation,
- stability, reflecting the consistency of feature ranking and behavior across model variants.

Each component was normalized within feature space using min–max scaling. The FRS was defined as the mean of these three feature-level components and was subsequently re-normalized within each variant.

In addition, two intermediate constructs were defined:

- stability score, combining relative rank position and cross-variant consistency,
- functional consistency score, combining ablation magnitude and top-*k* overlap between attribution and ablation.

The best-model diagnostic reliability score derived from permutation-based model diagnostics was preserved as contextual variant-level information, but was not included directly in the final feature-level FRS.

### Cross-variant consistency

To assess whether feature behavior was consistent across feature spaces, a cross-variant consistency metric was computed for features appearing in multiple variants. For each shared feature, attribution values were normalized within variants, and the spread across variants was quantified. Features with lower cross-variant spread were assigned higher consistency scores.

For features present in only a single feature space, cross-variant consistency was treated as not estimable rather than artificially maximal.

### Feature classification

Features were categorized into publication-oriented interpretability classes based on normalized attribution, functional impact, and stability scores. The following heuristic thresholds were applied:

- TRUE_SIGNAL: high attribution, high functional impact, and high stability,
- SPURIOUS_IMPORTANCE: high attribution but low functional impact,
- HIDDEN_SIGNAL: low attribution but high functional impact,
- LOW_SIGNAL: low across the evaluated dimensions,
- UNRESOLVED: incomplete feature-level component coverage preventing full classification.

Thresholds were defined at 0.67 on a normalized scale and were used for stratification rather than formal statistical inference. To evaluate the robustness of this heuristic choice, a dedicated threshold-sensitivity analysis was performed using alternative cutoffs (0.50, 0.60, 0.67, 0.75, and 0.80).

### Contradiction analysis

To explicitly quantify discrepancies between permutation-based attribution and functional importance, a contradiction framework was developed. For each feature, the difference between normalized attribution and ablation impact was computed as:

- signed contradiction: importance score minus ablation score,
- absolute contradiction: magnitude of disagreement.

Contradiction analysis was conducted primarily on complete-case features, defined as features with available attribution, ablation, stability, and FRS components. Features lacking sufficient information for complete contradiction profiling were retained in the full tables and classified as UNRESOLVED.

Features were further classified into contradiction categories:

- CONVERGENT_SIGNAL: high attribution, high functional impact, high reliability, and high stability,
- ATTRIBUTION_EXCESS: high attribution but low functional impact,
- LATENT_DEPENDENCE: low attribution but high functional impact,
- RELIABLE_NONDOMINANT: high reliability and stability despite limited attribution magnitude,
- LOW_EVIDENCE: complete-case features with low support across dimensions,
- UNRESOLVED: incomplete component coverage precluding contradiction classification.

These categories were used to characterize interpretability failure modes and to identify features whose apparent importance diverged from their functional contribution to model performance.

### Threshold sensitivity and unresolved-feature audit

Because the interpretability classes were defined using heuristic thresholds, a dedicated threshold-sensitivity analysis was performed to assess the robustness of feature classification across alternative cutoffs. Agreement with the baseline threshold (0.67) and stability of top-ranked features across thresholds were quantified within each feature space.

In addition, an unresolved-feature audit was conducted to determine the extent and origin of incomplete feature-level classification. This audit summarized the proportion of unresolved features in each variant and identified whether unresolved status was attributable to missing attribution components, missing ablation components, missing stability components, or multiple simultaneous missing components.

These supplementary analyses were used to distinguish genuine low-evidence signals from limitations in interpretability coverage.

### Statistical analysis

Associations between attribution magnitude, functional impact, and composite reliability were assessed using Spearman’s rank correlation coefficients within each feature space.

Overlap between top-ranked features based on attribution and ablation was quantified using top-*k* intersection analysis. In addition, the magnitude of disagreement between attribution and ablation was summarized using mean and median absolute contradiction scores.

All metrics were computed independently within each feature space to avoid cross-contamination of distributions.

### Software and reproducibility

All analyses were conducted using Python, with data processing performed using pandas and numerical operations using NumPy. Visualization was performed using Matplotlib.

The analytical workflow was organized into modular scripts (S1–S18), ensuring reproducibility of the full pipeline. Intermediate outputs from each stage were stored as structured CSV files and used as inputs for downstream analyses. Configuration snapshots were saved for key interpretability stages to document threshold choices and analysis settings.

## Results

### Attribution magnitude does not reflect functional importance

Across all feature spaces, permutation-based attribution magnitude did not align with functional importance functional importance measured through feature ablation. Instead, a systematic discrepancy between these two dimensions was consistently observed (Figure 1).

**Figure 1.**
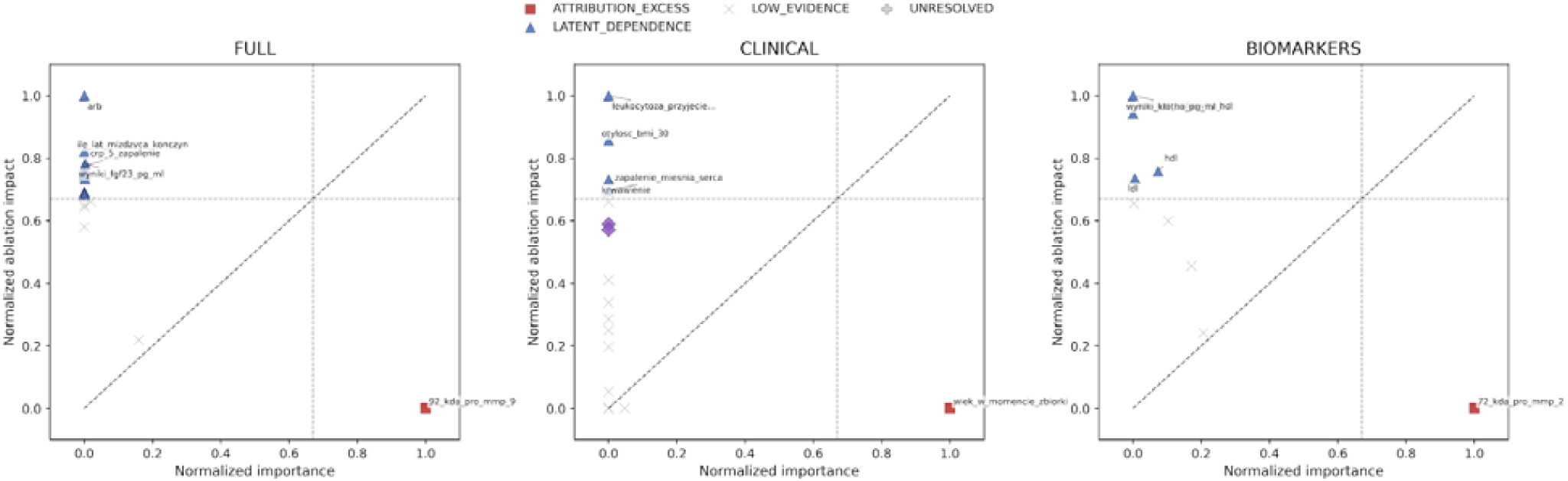
Attribution-based importance versus functional dependence across contradiction classes

Spearman correlations between permutation importance and ablation-derived performance change were weak or negative across all variants (Table 1). This effect was most pronounced in the BIOMARKERS feature space (ρ = −0.917), indicating a strong inverse relationship between attribution magnitude and functional dependence. Negative associations were also observed in the CLINICAL (ρ = −0.509) and FULL (ρ = −0.374) variants.

**Table 1.** Variant-level correlation and contradiction summary.

| **Variant** | **FULL** | **CLINICAL** | **BIOMARKERS** |
| --- | --- | --- | --- |
| Number of features | 54 | 43 | 10 |
| Spearman rho: importance vs ablation | -0.374 | -0.383 | -0.929 |
| Spearman rho: importance vs FRS | 0.53 | 0.453 | 0.816 |
| Spearman rho: ablation vs FRS | 0.259 | 0.62 | -0.533 |
| Top-k overlap: importance vs ablation | 1 | 0 | 1 |
| Mean absolute contradiction | 0.713 | 0.384 | 0.648 |
| Median absolute contradiction | 0.745 | 0.313 | 0.685 |
| Attribution excess features, n | 1 | 1 | 1 |
| Latent dependence features, n | 14 | 4 | 4 |
| Reliable high-impact features, n | 1 | 2 | 1 |
| Complete-case features, n | 20 | 20 | 9 |
| Unresolved features, n | 34 | 23 | 1 |

These results demonstrate that features ranked highly by permutation importance are not necessarily those on which model performance most strongly depends. Notably, several features with near-zero attribution produced substantial performance degradation when removed, indicating that attribution magnitude alone fails to capture functional relevance.

Collectively, these results demonstrate that attribution-based rankings fail to consistently identify functionally critical features, highlighting a systematic disconnect between attribution magnitude and model dependence.

### Systematic contradictions between attribution and model dependence

To quantify discrepancies between attribution and functional importance, a contradiction score was defined as the difference between normalized permutation importance and normalized ablation impact.

The distribution of signed contradiction scores revealed distinct patterns across feature spaces (Figure 2). In the FULL and BIOMARKERS variants, contradiction scores were predominantly negative, indicating that many features with low attribution exhibited substantial functional impact. In contrast, the CLINICAL variant showed a broader and more symmetric distribution, reflecting more heterogeneous relationships between attribution and dependence.

**Figure 2.**
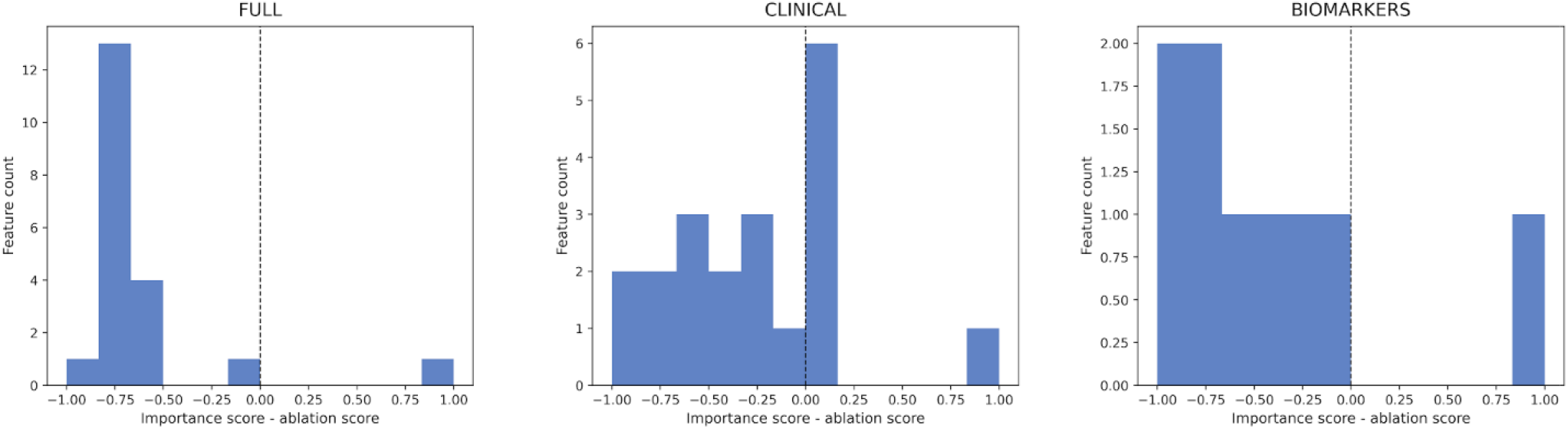
Distribution of signed interpretability contradictions

At the feature level, most observations deviated substantially from the identity line (importance = functional impact) (Figure 1), confirming that attribution magnitude and functional dependence represent distinct and frequently conflicting dimensions of model behavior.

### Distinct interpretability profiles across feature spaces

The composition of contradiction classes varied markedly between feature spaces, demonstrating that interpretability is strongly dependent on feature space definition (Figure 3).

**Figure 3.**
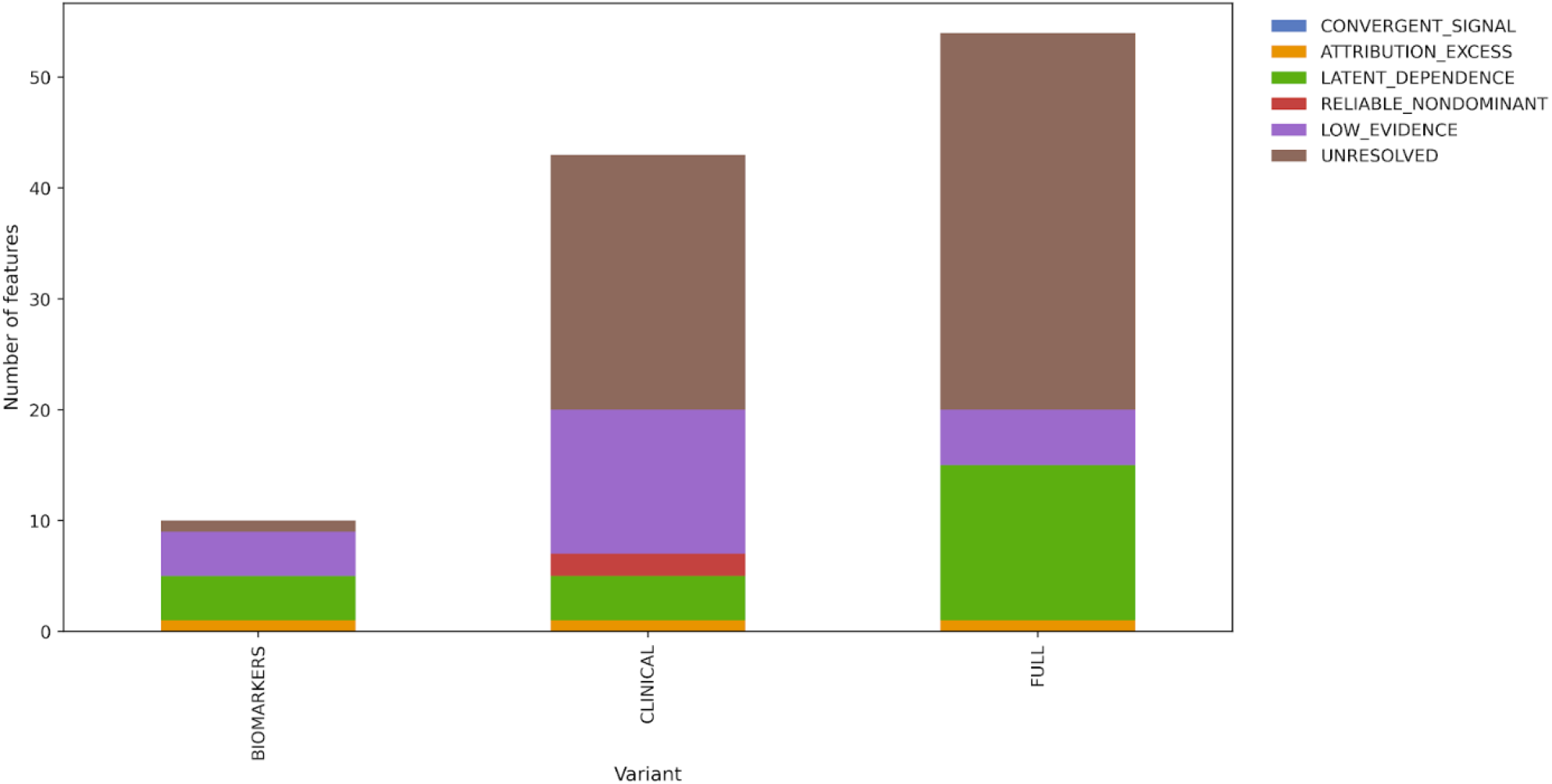
Contradiction-class composition across feature spaces

The FULL feature space was characterized by a predominance of features exhibiting latent dependence, indicating a large number of variables with low attribution but high functional impact. This pattern is consistent with redundancy and collinearity in combined feature sets, where predictive signals may be distributed across correlated variables.

The CLINICAL feature space exhibited a more heterogeneous profile, including both attribution excess and a predominance of features exhibiting latent dependence patterns. This suggests that clinical variables contain both genuinely informative and potentially misleading signals, depending on their role within the model.

In the BIOMARKERS feature space, a clear separation between attribution-driven and functionally important features was observed. Several biomarkers with high attribution showed limited functional impact, whereas others with modest attribution contributed substantially to model performance.

Across all feature spaces, convergent signal features were rare, indicating that few features simultaneously exhibited high attribution, high functional impact, and high stability.

These findings demonstrate that interpretability is not an intrinsic property of individual features, but rather depends on the structure and composition of the feature space.

### Identification of attribution excess and latent dependence features

The contradiction framework enabled the identification of two primary classes of interpretability failure (Figure 4).

**Figure 4.**
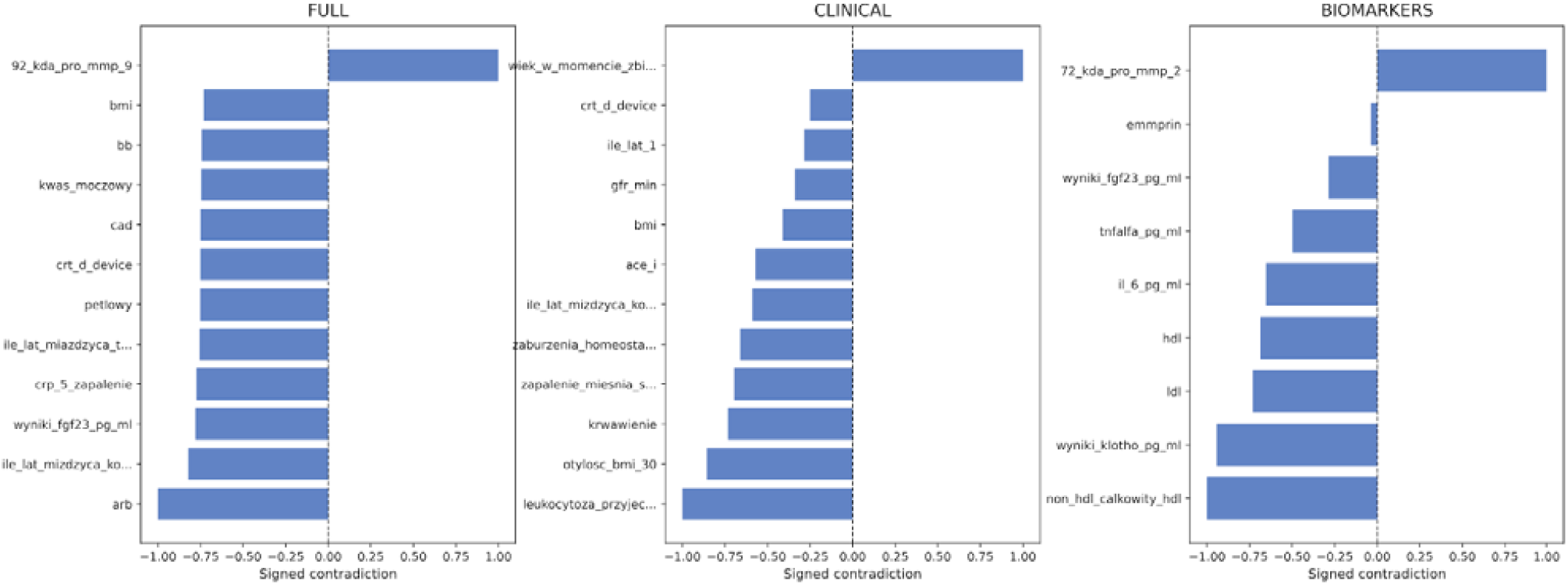
Top contradiction features by variant

Features exhibiting attribution excess, characterized by high permutation importance but low functional impact, represent variables that appear influential but do not materially contribute to predictive performance. These features likely reflect surrogate signals, correlations, or model-specific artifacts.

Latent dependence features, characterized by low attribution but high functional impact, represent variables that are essential for model performance despite being underestimated by attribution methods. These features are particularly important, as they would be overlooked by attribution-based feature selection strategies.

Latent dependence features were especially prominent in the FULL and BIOMARKERS variants, whereas attribution excess features were present but comparatively less frequent.

### Composite feature reliability reveals stable but non-dominant signals

Integration of attribution magnitude, functional impact, and stability into the Feature Reliability Score (FRS) enabled the identification of features that are consistently informative across multiple dimensions (Figure 5).

**Figure 5.**
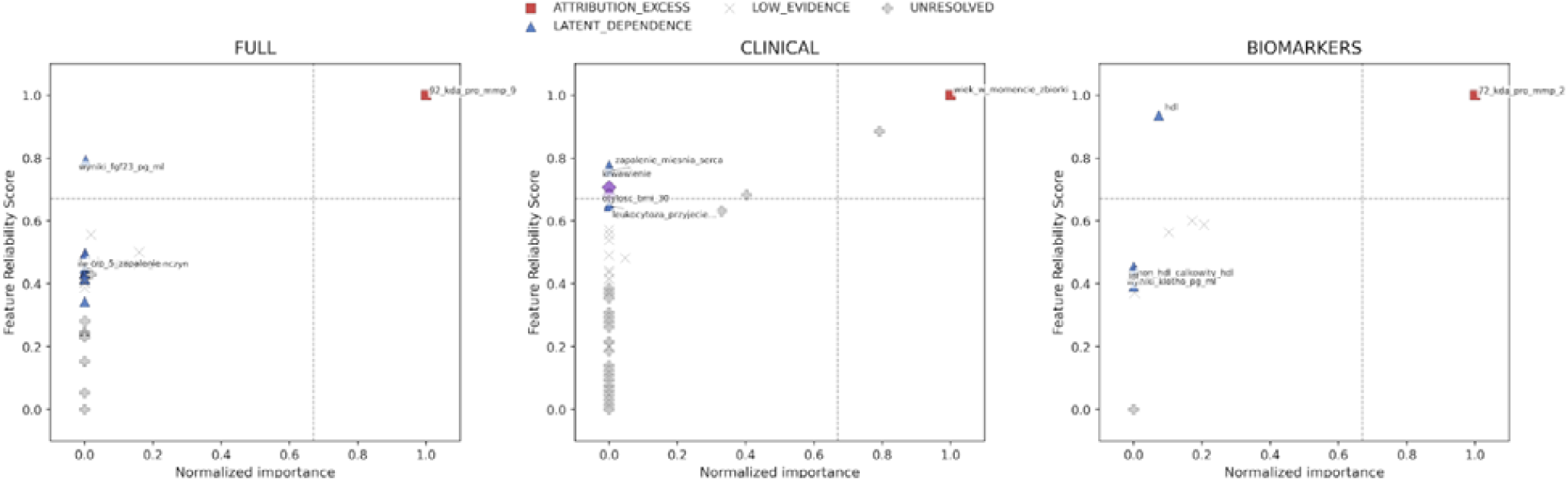
Importance versus composite reliability by contradiction class

Features with high FRS were not necessarily those with the highest attribution magnitude. Instead, reliable features typically exhibited moderate attribution combined with high stability and substantial functional impact.

This indicates that robust feature interpretation requires a multi-dimensional perspective. Features that are stable across perturbations and associated with measurable performance degradation provide more reliable evidence of model dependence than features than features identified solely based on attribution magnitude.

### Minimal overlap between attribution-based and function-based feature rankings

Comparison of top-ranked features based on permutation importance and ablation impact revealed minimal overlap across feature spaces (Table 1). In some cases, no shared features were identified between the top-*k* sets defined by these two criteria.

This lack of overlap further supports the conclusion that attribution-based rankings do not reliably identify functionally important features.

### Interpretability is constrained by feature-space coverage

A substantial proportion of features in the FULL and CLINICAL variants could not be fully classified due to incomplete component availability and were therefore labeled as UNRESOLVED. In contrast, the BIOMARKERS feature space exhibited a substantially lower proportion of unresolved features.

This indicates that interpretability is not only a function of model behavior, but also of feature-space completeness and analytical coverage. Larger and more heterogeneous feature spaces may introduce structural limitations that reduce interpretability, even when predictive performance is high.

### Interpretability as stability under perturbation

Taken together, these results demonstrate that feature attribution based on distributional perturbation does not provide a complete or stable representation of model behavior.

Instead, functional importance derived from structural perturbation (feature removal), combined with stability and consistency across feature spaces, provides a more robust characterization of feature relevance.

These findings support a reinterpretation of model interpretability as a property of stability under perturbation, rather than as a direct consequence of attribution magnitude.

## Discussion

### Interpretability as a multi-dimensional and unstable property

In this study, we demonstrate that widely used feature attribution methods do not provide a reliable or complete representation of model behavior. Across multiple feature spaces, attribution magnitude derived from permutation importance was systematically misaligned with functional importance assessed through feature ablation. These discrepancies were not incidental but consistent, reproducible, and structurally dependent on feature space composition.

Our findings challenge the implicit assumption that attribution magnitude can serve as a proxy for feature relevance. Instead, they indicate that interpretability is inherently multi-dimensional and cannot be captured by a single metric. In particular, attribution, functional dependence, stability, and model-level reliability represent partially independent dimensions that may diverge substantially. This perspective aligns with a growing body of work highlighting the instability, non-uniqueness, and context dependence of post hoc explanation methods [11–13,18–21].

### Limitations of attribution-based explanations

Permutation-based attribution methods quantify the sensitivity of model predictions to distributional perturbations. However, our results demonstrate that this form of perturbation does not necessarily reflect the structural role of a feature within the model [7,8,18].

Features identified as highly important by permutation-based methods were frequently found to have a limited impact on model performance when removed. These attribution excess features may arise from correlations, surrogate relationships, or higher-order interactions rather than direct functional contributions [6,9,22].

Conversely, features with low attribution but high ablation impact - defined here as features exhibiting latent dependence a critical blind spot of attribution-based approaches. Such features would be overlooked in standard feature selection pipelines, despite being essential for maintaining model performance. This observation is consistent with prior studies demonstrating that feature importance measures can fail in the presence of correlated or redundant predictors [6,15,22,23].

Taken together, these results indicate that attribution-based explanations alone are insufficient for identifying functionally relevant features in complex predictive models, particularly in high-dimensional biomedical settings.

### Interpretability depends on the feature space structure

A key finding of this study is that interpretability is not an intrinsic property of individual features but emerges from the structure of the feature space. The FULL, CLINICAL, and BIOMARKERS variants exhibited distinct contradiction profiles, with different balances between attribution excess, latent dependence, and convergent signals.

The dominance of latent dependence features in the FULL feature space suggests that redundancy and collinearity can obscure attribution signals. When multiple correlated variables encode similar information, permutation-based methods may distribute importance across features in a way that underestimates their individual functional contributions. This phenomenon is well documented in studies of model reliance and feature dependence [6,15,23].

In contrast, the CLINICAL feature space showed a more heterogeneous structure, including both convergent and misleading signals, reflecting the complex interplay between routinely collected variables. The BIOMARKERS feature space demonstrated a clear separation between attribution-driven and functionally important features, indicating that even in molecular data, attribution may fail to identify the most critical predictors.

These findings emphasize that interpretability analyses must be contextualized within the feature space under consideration, rather than treated as universally applicable. This is particularly important in biomedical applications, where feature redundancy, measurement noise, and heterogeneous data structures are common [1,2,16].

### Stability under perturbation as a criterion for interpretability

The integration of attribution, ablation, and stability into a unified framework revealed that reliable features are not necessarily those with the highest attribution magnitude. Instead, features that consistently contribute to model performance across perturbations and analytical contexts provide stronger evidence of relevance.

This observation supports a reinterpretation of interpretability as stability under perturbation. In this view, a feature is considered interpretable not because it has a high attribution score, but because its relevance is robust to changes in data distribution, model specification, and feature space composition. This perspective is increasingly emphasized in recent work on trustworthy and reliable AI systems [19–21,24].

The Feature Reliability Score introduced in this study operationalizes this concept by integrating complementary dimensions of feature behavior. While the specific formulation of this score is heuristic, it demonstrates the value of combining attribution, functional impact, and stability to obtain a more robust assessment of feature relevance.

### Implications for feature selection and model interpretation

The discrepancies identified in this study have important implications for both methodological practice and applied modeling.

First, reliance on attribution-based rankings for feature selection may lead to suboptimal or misleading conclusions. Features that appear highly important may not contribute meaningfully to model performance, while functionally critical features may be excluded due to low attribution scores.

Second, interpretability assessments should incorporate multiple forms of perturbation. Distributional perturbations (e.g., permutation) capture the sensitivity of predictions, whereas structural perturbations (e.g., feature removal) capture model dependence. Both perspectives are necessary for a comprehensive understanding of model behavior.

Third, stability across analytical contexts—including feature spaces, model configurations, and perturbation strategies—should be treated as a central criterion for interpretability, particularly in high-dimensional biomedical applications where model reliability is critical [1,17,24].

### Limitations

Several limitations should be considered when interpreting these findings.

First, the thresholds used for feature classification were heuristic and intended for stratification rather than formal inference. Although sensitivity analysis (S17) confirmed that the main conclusions are robust to threshold variation, alternative thresholding strategies may yield different class boundaries.

Second, some features could not be evaluated through ablation due to missing or undefined performance estimates and were therefore classified as unresolved. While these cases were retained to reflect the full analytical pipeline, introduce some uncertainty into feature-level interpretation.

Third, the Feature Reliability Score was intentionally defined as a feature-level construct and does not directly incorporate model-level reliability into the composite score. While this design improves interpretability, it may omit relevant global context.

Finally, the observed discrepancies may be influenced by feature redundancy and collinearity, particularly in the FULL feature space. Although this reflects realistic data conditions, it complicates attribution of importance to individual variables.

## Conclusions

This study demonstrates that feature attribution methods alone are insufficient for reliable model interpretation. By integrating attribution, functional impact (defined as performance degradation upon feature removal), and stability into a unified framework, we show that interpretability is inherently multi-dimensional, context-dependent, and cannot be reliably inferred from attribution magnitude alone.

Beyond these findings, this work introduces three conceptual and methodological advances: (i) a systematic quantification of disagreement between attribution and functional importance, (ii) a formalization of interpretability as stability under perturbation, and (iii) a composite reliability framework for feature evaluation.

Our results support a shift from attribution-centric explanations toward frameworks that explicitly account for stability under perturbation. Such approaches provide a more robust and informative basis for understanding model behavior, particularly in high-dimensional and correlated biomedical data settings.

More broadly, these findings highlight the need for multi-dimensional interpretability frameworks that move beyond single-metric explanations and enable more reliable, reproducible, and clinically meaningful insights from machine learning models.

## Data Availability

The data that support the findings of this study are not publicly available due to privacy and ethical restrictions but are available from the corresponding author upon reasonable request. The analytical code used in this study is publicly available at: https://github.com/npiorkowska-science/ml-interpretability-stability-framework

## Data and Code Availability

The analytical code used in this study is publicly available in a dedicated GitHub repository: https://github.com/npiorkowska-science/ml-interpretability-stability-framework

## Funding

This research received no specific grant from any funding agency in the public, commercial, or not-for-profit sectors.

## Conflict of Interest

The authors declare that they have no conflicts of interest relevant to this study.

## Ethics Statement

The study was conducted in accordance with the Declaration of Helsinki. The use of clinical data for the proof-of-concept analysis was approved by the Ethics Committee of the Medical University of Wroclaw (approval numbers: KB-54/2019, KB-514/2019, KB-387/2021). All data used in the analysis were anonymized prior to processing. The systematic review part of the study was conducted using previously published studies and did not require additional ethical approval.

## Author Contributions

Natalia Piórkowska conceptualized the study, designed the methodology, performed the analysis, interpreted the results, and wrote the manuscript.

Agnieszka Olejnik led the acquisition and curation of patient clinical and biomarker data, performed laboratory and diagnostic analyses, ensured data integrity and resource management, and contributed to data interpretation and critical revision of the manuscript.

Alan Ostromęcki conceptualized the study, designed the methodology, visualization and wrote the manuscript.

Wiktor Kuliczkowski led the acquisition and curation of patient clinical and biomarker data.

Andrzej Mysiak led the acquisition and curation of patient clinical and biomarker data.

Iwona Bil-Lula led the acquisition and curation of patient clinical and biomarker data, performed laboratory and diagnostic analyses, language correction and contributed to data interpretation and critical revision of the manuscript.

## Acknowledgments

The authors acknowledge the Wroclaw Medical University for providing access to the clinical data used in the proof-of-concept analysis. The authors also thank collaborators who contributed to the development of the methodological framework and provided valuable feedback during the preparation of this study.

## Notes

### Competing Interest Statement

The authors have declared no competing interest.

### Author Declarations

Ethics Committee of the Medical University of Wroclaw granted ethical approval for this study. The use of clinical data was approved by the Ethics Committee of the Medical University of Wroclaw. All data were anonymized prior to analysis.

## References

1. Esteva, A. et al. A guide to deep learning in healthcare. Nat. Med. 25, 24–29 (2019).

2. Rajkomar, A., Dean, J. & Kohane, I. Machine learning in medicine. N. Engl. J. Med. 380, 1347–1358 (2019).

3. Topol, E. Deep Medicine: How Artificial Intelligence Can Make Healthcare Human Again. Basic Books (2019).

4. Lundberg, S. M. & Lee, S.-I. A unified approach to interpreting model predictions. Adv. Neural Inf. Process. Syst. 30 (2017).

5. Breiman, L. Random forests. Mach. Learn. 45, 5–32 (2001).

6. Fisher, A., Rudin, C. & Dominici, F. All models are wrong, but many are useful: learning a variable’s importance by studying an entire class of prediction models. J. Mach. Learn. Res. 20, 1–81 (2019).

7. Hooker, S., Erhan, D.Kindermans, P.-J. & Kim, B. A benchmark for interpretability methods in deep neural networks. Adv. Neural Inf. Process. Syst. (2019).

8. Covert, I., Lundberg, S. & Lee, S.-I. Understanding global feature contributions with additive importance measures. Adv. Neural Inf. Process. Syst. (2020).

9. Molnar, C. Interpretable Machine Learning. 2nd ed. (2022).

10. Rudin, C. Stop explaining black box machine learning models for high stakes decisions and use interpretable models instead. Nat. Mach. Intell. 1, 206–215 (2019).

11. Adebayo, J. et al. Sanity checks for saliency maps. Adv. Neural Inf. Process. Syst. (2018).

12. Ghorbani, A., Abid, A. & Zou, J. Interpretation of neural networks is fragile. AAAI (2019).

13. Slack, D. et al. Fooling LIME and SHAP: adversarial attacks on post hoc explanation methods. AIES (2020).

14. Hooker, S. et al. What do compressed deep neural networks forget? Int. Conf. Learn. Represent. (ICLR) (2020).

15. Li, J., Rudin, C. & others. Measuring feature importance through model reliance. J. Mach. Learn. Res. 23 (2022).

16. Riley, R. D. et al. Minimum sample size for developing a multivariable prediction model. Stat. Med. 39, 1262–1275 (2020).

17. Collins, G. S. & Moons, K. G. Reporting of artificial intelligence prediction models. BMJ 368, m689 (2020).

18. Krishna, S., Han, T., Gu, A. & Wu, Z. The disagreement problem in explainable AI: a practitioner’s perspective. Proc. ACM Conf. Fairness, Accountability, and Transparency (FAccT) (2022).

19. Doshi-Velez, F. & Kim, B. Towards a rigorous science of interpretable machine learning. arXiv:1702.08608 (updated discussions 2021).

20. Amparore, E. et al. Feature important explanations under correlation: challenges and limitations of SHAP. Sci. Rep. 11, 1–13 (2021).

21. Frye, C., Rowat, C. & Feige, I. Asymmetric feature dependence in machine learning models. Adv. Neural Inf. Process. Syst. (2020).

22. Watson, D. et al. On the robustness of feature attribution methods. Proc. ICML (2022).

23. Carvalho, D. V., Pereira, E. M. & Cardoso, J. S. Machine learning interpretability: a survey on methods and metrics. Electronics 10, 832 (2021).

